# Patient and caregiver perceptions of electronic health records interoperability in the NHS and its impact on care quality: A focus group study

**DOI:** 10.1101/2024.01.30.24302031

**Authors:** Edmond Li, Olivia Lounsbury, Jonathan Clarke, Hutan Ashrafian, Ara Darzi, Ana Luisa Neves

## Abstract

**Background:** The proliferation of electronic health records (EHR) in health systems of many high-income countries has ushered in profound changes to how clinical information is used, stored, and disseminated. For patients, being able to easily access and share their health information electronically through interoperable EHRs can often impact safety and their experience when seeking care across healthcare providers. While extensive research exists examining how EHRs affected workflow and technical challenges such as limited interoperability, much of it was done from the viewpoint of healthcare staff rather than from patients themselves. This leaves a critical knowledge gap in our evidence base to inform better implementation of health information technologies which needs addressing.

**Aims and Objectives:** This study aimed to explore how patients with chronic conditions or polypharmacy and their caregivers perceive the current state of EHR interoperability, identify instances where it was associated with negative health outcomes, and elucidate patient-driven recommendations to address concerns raised.

**Methods:** A total of 18 patients and caregivers participated in five online focus groups between May-July 2022. Thematic analysis was performed to generate codes and derive higher-order themes.

**Results:** Participants highlighted that EHR interoperability in the NHS does not meet patient needs and expectations. While patients’ understanding of the concept of EHR interoperability was mixed, most were able to describe how the inability to seamlessly share health information within EHR has negatively impacted care. Limited interoperability contributed to inaccurate medical records, perpetuated existing incorrect information, impaired clinical decision-making, and often required patients to resort to using workarounds. Patients also voiced ideas for potential solutions for consideration. These included a move towards a one-centralised system approach, strengthening data security measures to augment other efforts to increase interoperability, prioritising health information technology training for NHS staff, and involving more allied health professionals and patients themselves in the EHR data curation process.

**Conclusion:** Our study contributes to the existing body of literature by providing the perspectives of patients and carers most likely to encounter interoperability challenges and therefore those most ideally positioned to propose potential solutions. As highlighted by patients, researchers and policymakers should consider social, educational, and organisational solutions, in addition to technical solutions.

**Public Interest Summary:** Lack of interoperability, *i.e.,* the ability to share a patient’s health information electronically between healthcare providers, can affect the quality of care received. However, much of the existing research was done from the viewpoint of healthcare staff rather than from patients themselves. This study explored the views of patients regarding what they feel interoperability in the NHS is currently like, how they think it affects their care, and what they think can be done to improve it. Patients reported interoperability to often be poor. It caused inconvenience both to themselves and their healthcare provider, and negatively impacted their experience getting care overall. Patient suggestions for improvement included centralising and reducing the number of existing systems, having more training for healthcare staff, and supporting patients and other healthcare staff in managing their health data.

## Introduction

Since the introduction of Electronic Health Records (EHR), research efforts focussed on exploring the benefits surrounding their implementation and use (1,2), impact on clinical workflows (3,4), provider experiences (5,6), as well as many of the limitations due to the lack of EHR interoperability (7,8).

In comparison, there is a relative paucity of evidence dedicated to investigating EHR interoperability from the perspective of patients or caregivers, and its impact on the perceived quality of care received. A study by *Chang et al.,* highlighted that patients who access care mostly within one health system and had all their medical records in one centralised EHR, tended to have a more positive impression of care coordination. This contrasted with patients who described a sense of *‘disconnect’* between their primary care provider and specialists when seeking care across two or more health systems using multiple non-interoperable EHR. The study concluded that patients *‘strongly endorsed the need for better communication, interoperable health records, and improved transitions of care between providers and health systems’* (9).

For patients living with chronic conditions or polypharmacy, their often-complex healthcare needs make them especially dependent on health information technologies, such as EHR, that support greater interoperability to ensure a safe delivery of care (9–11). As patient-centric care has become increasingly encouraged and patients are better empowered to adopt more assertive roles in their own healthcare decisions, a similar shift regarding the stewardship of healthcare data currently held in EHR can already be seen in some initiatives. For example, organisations such as *Understanding Patient Data* are already working with patient groups across the healthcare landscape to equip patients with the information needed to use their own data effectively (12). As patients become more empowered to use their data in their care, it is likely that their expectations of EHR will become increasingly voiced. Understanding patient and caregiver perspectives around interoperability, including their perceived risks and potential solutions, will help us anticipate these needs proactively in the design of new EHR systems and the development of healthcare data policies. Evidence gathered will be useful in informing future healthcare technology policies which should better reflect the bespoke needs of patients and caregivers and allow for the better leveraging of digital technologies integrated into the modern healthcare environment.

This study aims to investigate how patients and their caregivers perceive the current status and potential future of EHR interoperability in the NHS. Specific study objectives include:

a) To explore patients’ and caregivers’ knowledge, understanding, and expectations of concepts such as electronic health records and interoperability.
b) To explore their perception of the impact of interoperability on patient safety.
c) To explore potential solutions to address current EHR interoperability challenges.

## Methods

### Study Design

A qualitative methods approach using focus groups was selected for this study given that focus groups inherently amplify insights into particular topics as a result of the organic interactions and group dynamics between the participants themselves (13–15). Given the lack of extensive prior literature on the topic of EHR interoperability in the NHS from the patients’ perspective, focus groups are also well suited to explore participant perceptions and how they arrived at these conclusions (13). Thematic analysis of the focus group transcripts was performed by two members of the research team (EL, OL).

The Consolidated Criteria for Reporting Qualitative studies (COREQ) guidelines was used to ensure the findings in this report are aligned with best practices for reporting qualitative research. (16).

### Study Population

This study captured the views of adult patients and their caregivers who receive care at NHS facilities that make use of EHR systems. Patients with chronic conditions were chosen as this patient group is expected to experience the greatest reliance on the EHR due to their lengthy medical history, polypharmacy, and often-complex care needs spanning multiple healthcare providers.

The inclusion criteria for participants were as follows:

- Patients aged 18+ who have at least one chronic condition listed in the Charlson Comorbidity Index (17,18) or caregivers who provide care to patients with chronic conditions
- Patients or caregivers whose care frequently requires visits between at two or more healthcare facilities (*e.g.,* GP surgeries and hospitals)
- Able to communicate verbally in English
- Have internet access and equipment needed to perform video or telephone conferencing.

### Sampling and Recruitment

Participants were recruited using convenience sampling with the aim of creating four to five focus groups (19–23). Recruitment was completed in partnership with the *VOICE UK* patient involvement and engagement network in addition to using study advertisements posted on social media platforms (*e.g.,* Twitter, Facebook) (24). Participants did not have any relationship with the researcher beforehand and received no financial compensation for their participation.

### Data Collection

A topic guide containing nine open-ended questions exploring the three study aims was used **(Appendix 1)**. The topic guide was developed using evidence from existing available literature on the research topic and input from members of the [to be named after blinding// XXBLINDEDXX] Research Patients Group (RPG) Patient and Public Involvement and Engagement (PPIE) group. The topic guide was piloted with two PPIE groups and iteratively revised before its use.

Recruitment took place between May-June 2022. The focus groups were conducted between May-July 2022 and were facilitated in English via online conferencing platforms (*e.g.,* Zoom), with the option for participants to dial in via telephone. The sessions were digitally recorded for transcription verbatim by an independent transcription service. All study material was saved on password-secured servers at [to be named after blinding// XXBLINDEDXX]. Only members of the research team and participants were present at the focus groups. Participants did not review the transcripts or the findings. No follow-up focus groups were conducted.

### Data Analysis

Transcripts were coded and thematically analysed independently by two qualitative researchers with backgrounds in clinical medicine, public health, and patient safety (EL, OL). Regular meetings between members of the research team ensured coding quality and enabled iterative refinement of the codes and subthemes to form higher order themes. Overall, the analysis was both deductive (*i.e.,* partly based on pre-existing knowledge from the literature) and inductive (*i.e.,* additional concepts identified from the data derived from the transcripts) (25).

## Results

A total of 18 patients and caregivers participated in our study across five focus groups (minimum two, maximum six participants per group). The focus groups lasted on average 48 minutes per session.

The three main themes and respective subthemes are mapped in **Figure 1**. See **Table 1** for a breakdown of the participants’ basic demographic characteristics. Missing details on patient characteristics are due to unreturned demographic questionnaires.

**Figure 1:**
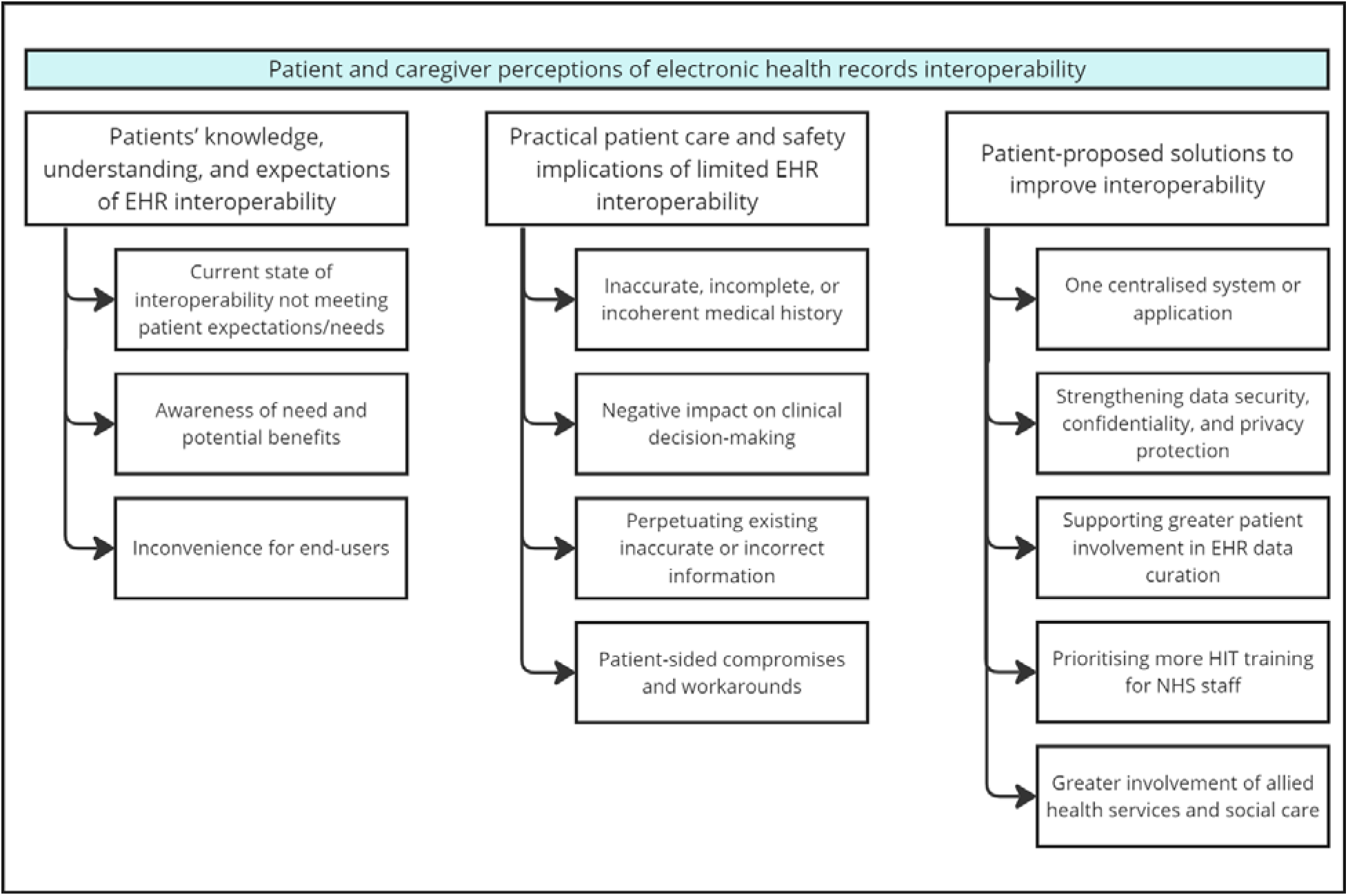
Mapping of prominent themes identified.

**Table 1:** Basic participant demographic characteristics.

| Characteristics |  | n (%) |
| --- | --- | --- |
| Gender | Male | 6 (33.33%) |
|  | Female | 11 (61.11%) |
|  | Other | 0 (0%) |
|  | Missing | 1 (5.56%) |
| Age | 18-24 years old | 1 (5.56%) |
|  | 25-34 years old | 0 (0%) |
|  | 35-44 years old | 2 (11.11%) |
|  | 45-54 years old | 2 (11.11%) |
|  | 55-64 years old | 6 (33.33%) |
|  | 65 years old and older | 6 (33.33%) |
|  | Missing | 1 (5.56%) |
| Ethnic Group | White (English/Welsh/Scottish/Northern Irish/British, Irish, Gypsy or Irish Traveller, Any other White background) | 11 (61.11%) |
|  | Mixed/Multiple Ethnic Groups (White and Black Caribbean, White and Black African, White and Asian, Any other Mixed/Multiple ethnic background) | 2 (11.11%) |
|  | Asian/Asian British (Indian, Pakistani, Bangladeshi, Chinese, Any other Asian background) | 2 (11.11%) |
|  | Black/African/Caribbean/Black British (African, Caribbean, Any other Black/African/Caribbean background) | 1 (5.56%) |
|  | Other Ethnic Group (Arab, Any other ethnic group) | 1 (5.56%) |
|  | Missing | 1 (5.56%) |

### Patients’ knowledge, understanding, and expectations of EHR interoperability

Patients demonstrated variable levels of understanding of EHR interoperability. While most were unfamiliar with the term, many participants deduced it pertained to how their clinical data was handled across the NHS. Participants associated the meaning of interoperability with having inadequate levels of access to their own clinical information (*i.e.,* via patient-facing online portals and mobile phone applications), or the lack of a more streamlined appointment scheduling experience.

A minority of patients who stated that they had had previous experience with digital healthcare technologies were better able to describe what EHR interoperability meant for them: the ability to access and share clinical data within connected EHR systems between various healthcare providers involved in their care. This definition was largely similar across this group of participants.

**Textbox 1:**
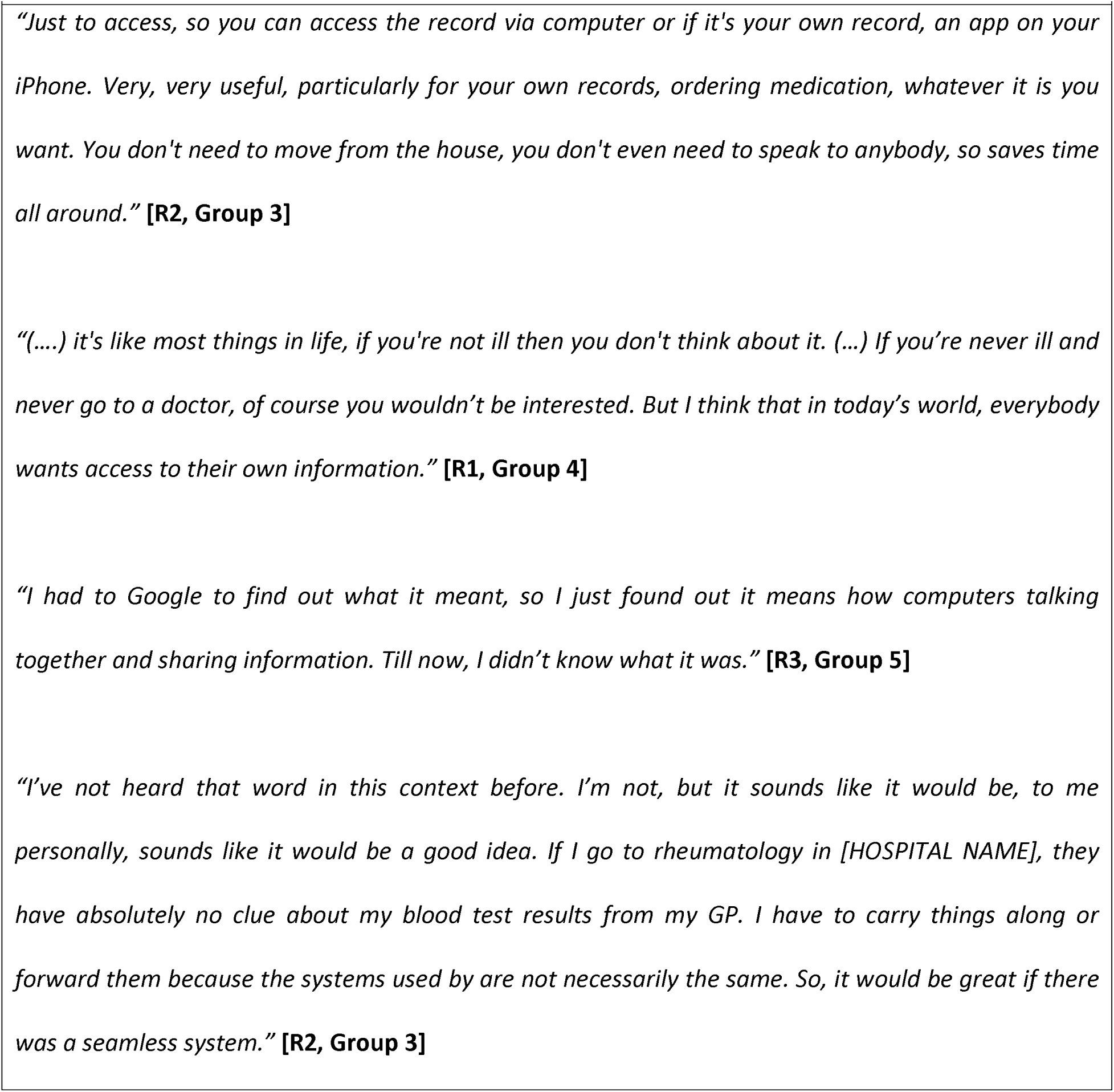
Patients’ knowledge, understanding, and expectations of EHR interoperability.

### Current state of interoperability not meeting patient expectations and needs

Participants largely agreed that the current implementation of EHR interoperability across the NHS is not adequately meeting their needs and expectations. Numerous participants detailed instances where their care was negatively affected due to the lack of interoperability (*e.g.,* need for repeat investigations, recounting of medication lists, documentation of incomplete or inaccurate medical histories, and miscommunications between their GP and specialists). These contributed to a frustrating, burdensome, and overall negative experience for patients and their caregivers when accessing both primary and secondary care.

**Textbox 2:**
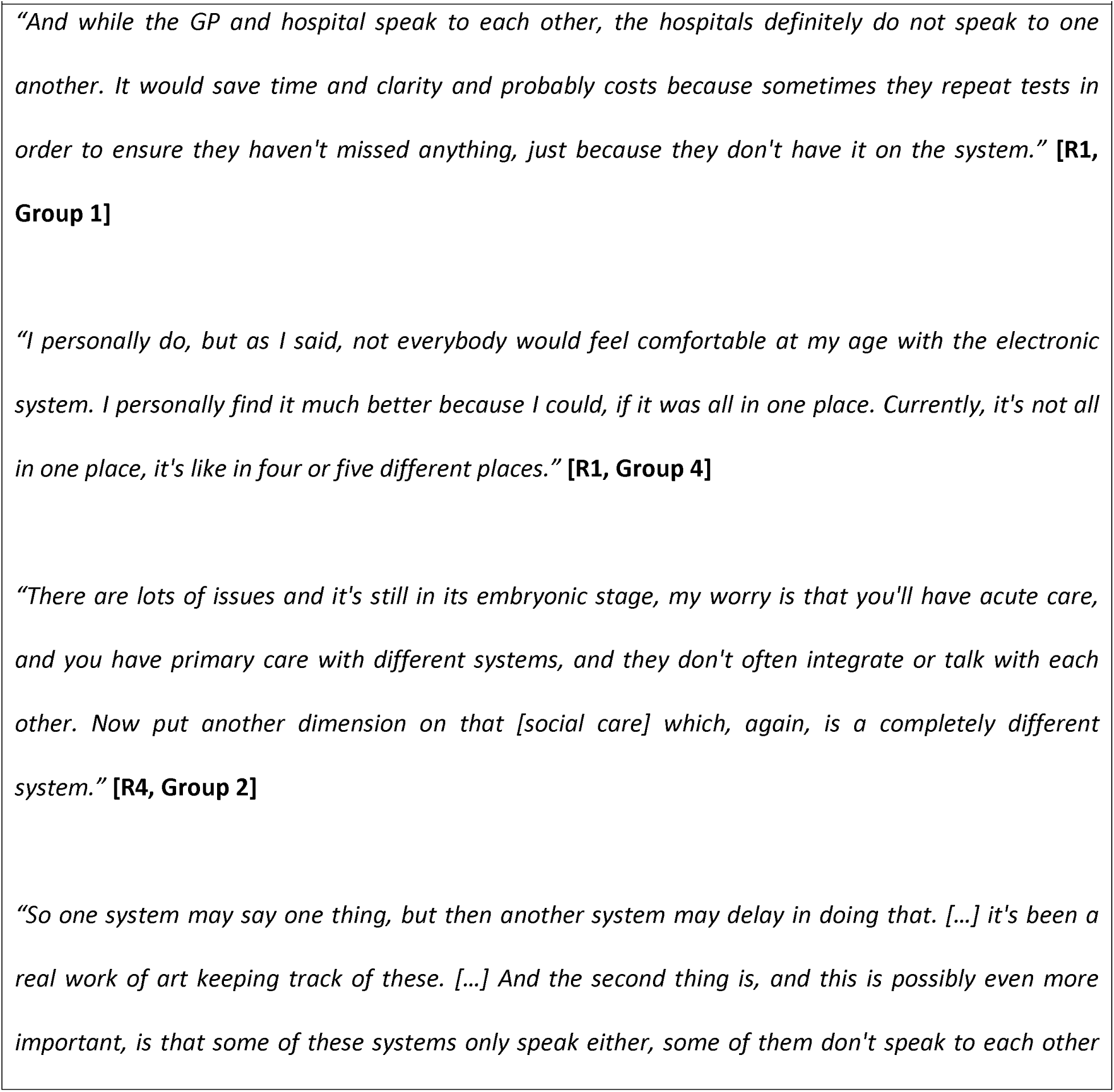
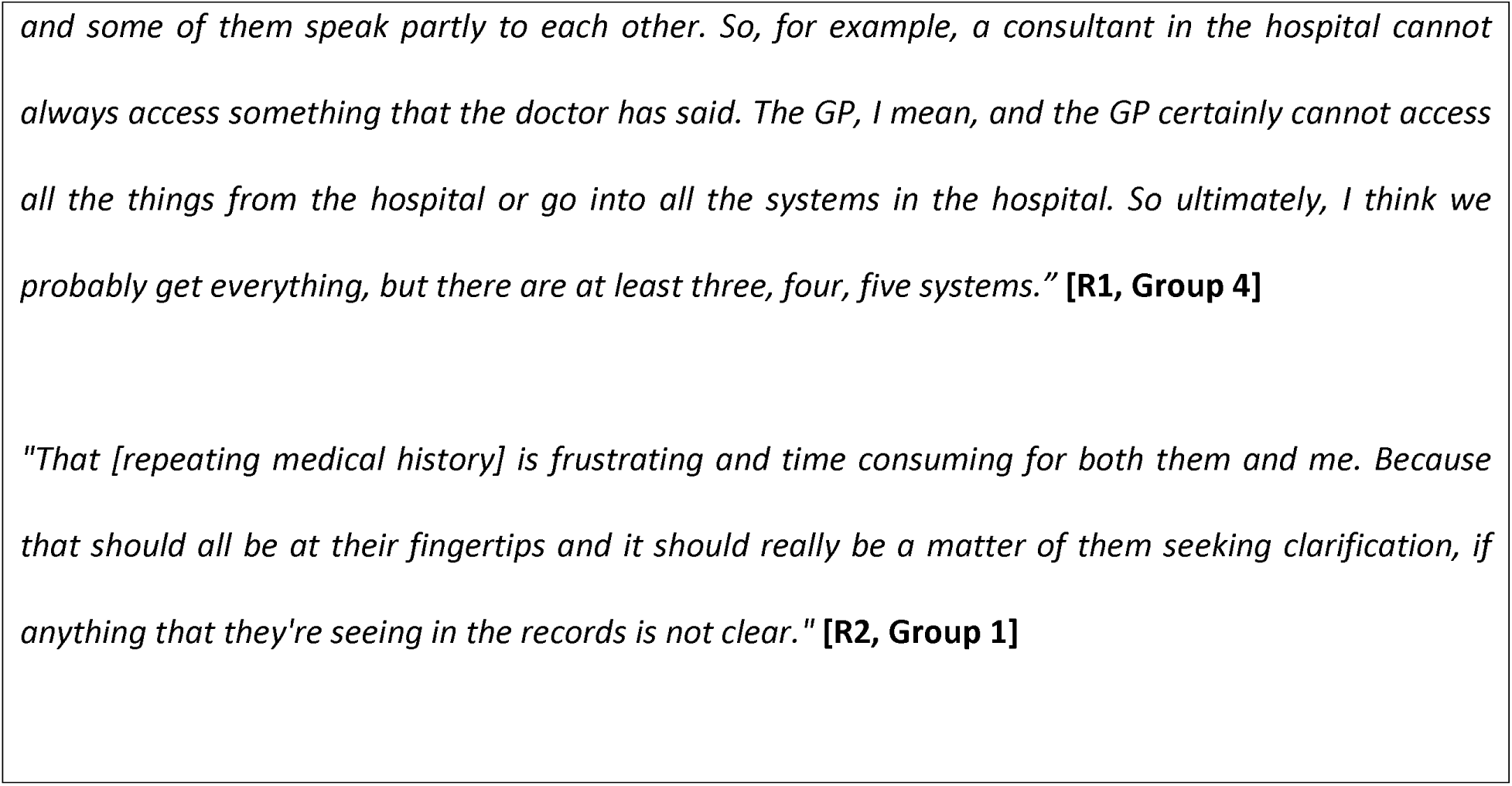
Current state of interoperability not meeting patient expectations/needs.

### Awareness of need and potential benefits

Participants were easily able to recognise the importance and potential benefits to their care of enhanced interoperability. Aside from the apparent, practical benefits such as convenience, interoperability was perceived to enhance patient advocacy and enable greater autonomy in decision-making and care planning. However, participants did indicate that the benefits of interoperability were contingent on the quality of data available in the EHR.

**Textbox 3:**
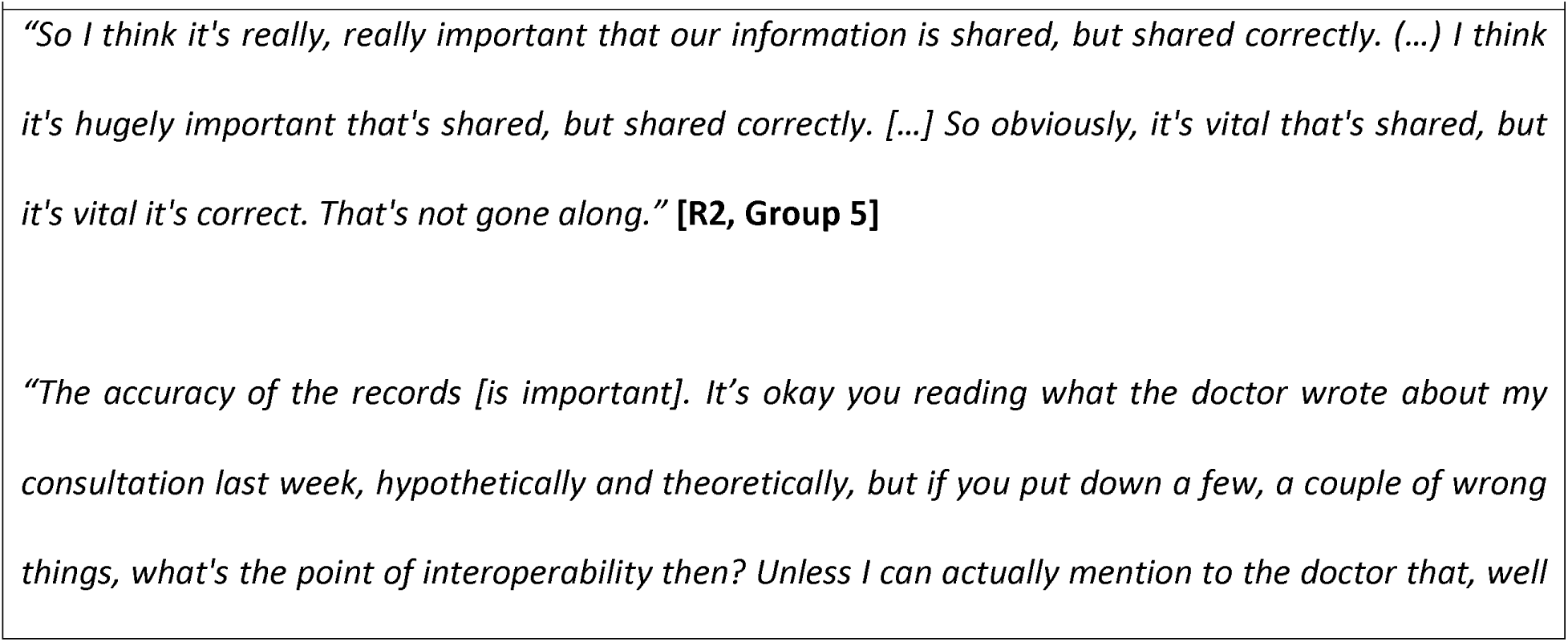
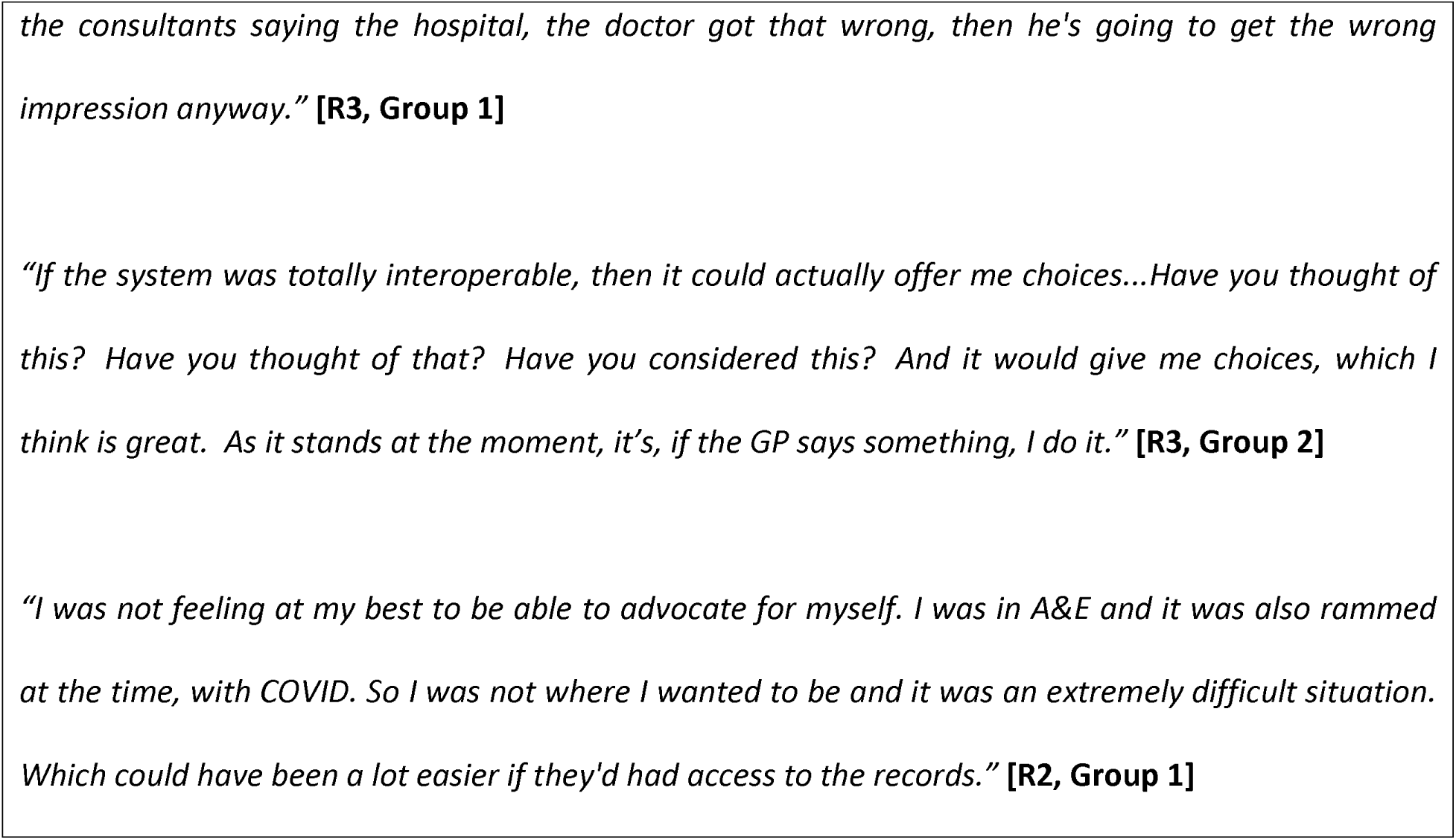
Awareness of need and potential benefits.

### Inconvenience for end-users

The greatest shortcoming of the current state of interoperability recognised by participants was the inconvenience caused, both to themselves and for healthcare providers. For patients, suboptimal interoperability was perceived to negatively impact continuity of care and pose safety risks as medical records were often incomplete and required rectification. Participants also perceived that inadequate interoperability often translated into suboptimal use of the limited clinical time with healthcare providers due to the amount of time spent reconciling patient information.

**Textbox 4:**
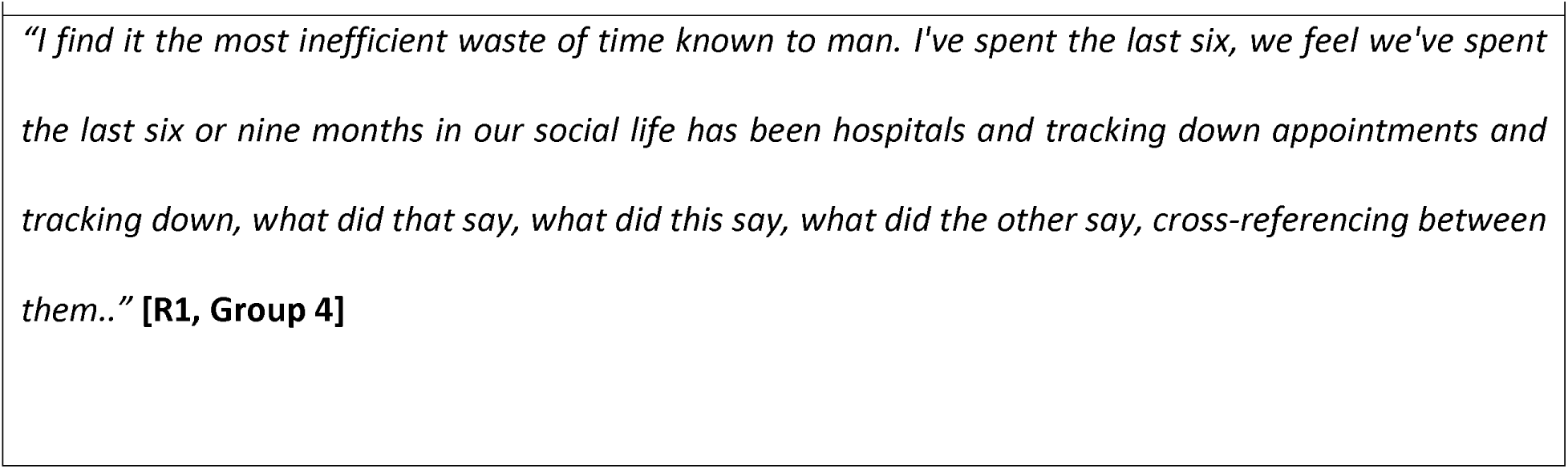
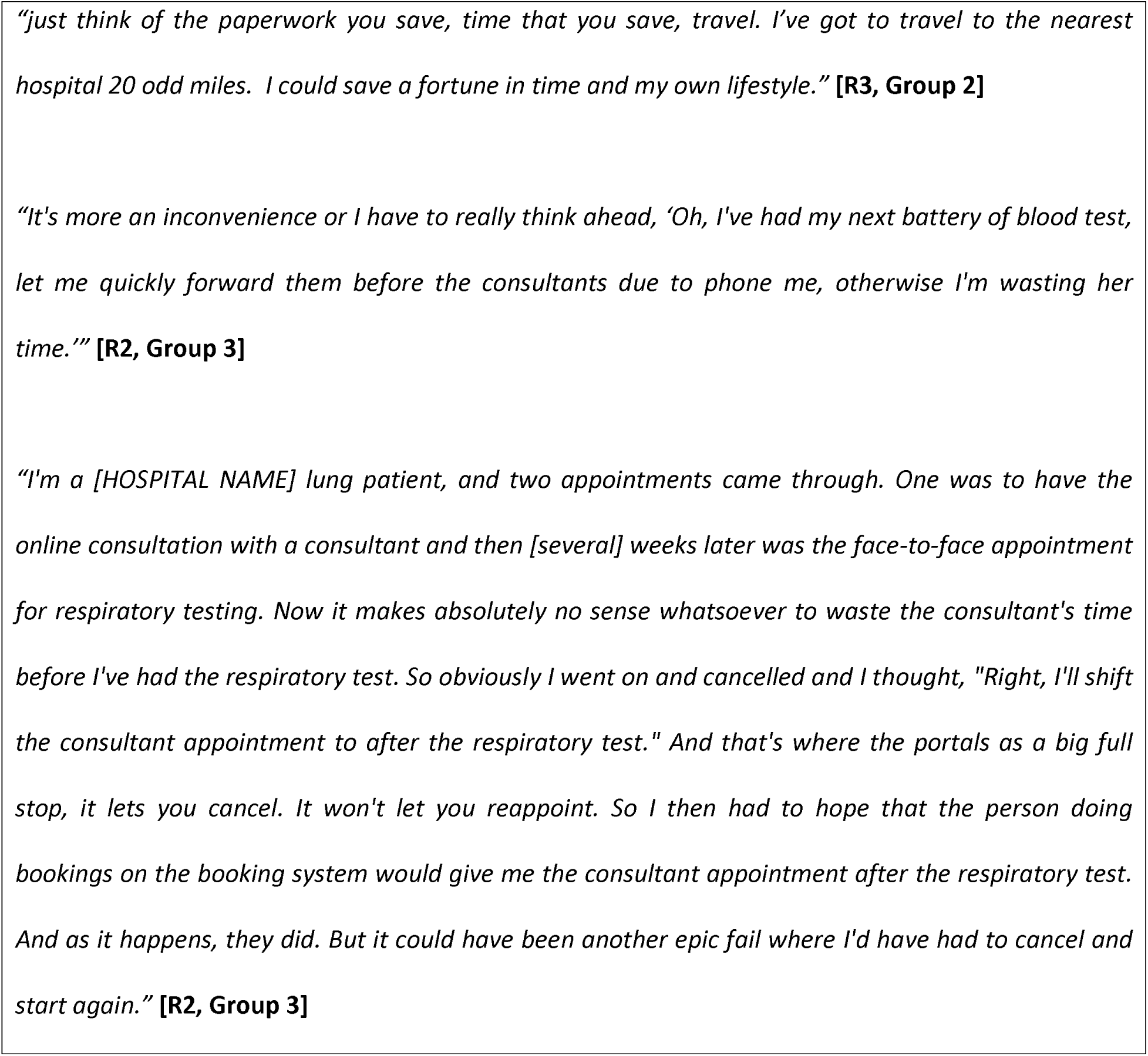
Inconvenience for end-users.

### Practical patient care and safety implications of limited EHR interoperability

Participants showed an extensive awareness of the direct, indirect, and potential impact posed to their immediate care and safety due to the general lack of EHR interoperability in the NHS.

### Inaccurate, incomplete, or incoherent medical history

Almost all participants described instances where they identified inaccurate or incomplete clinical information in their EHR during their clinical encounters, resulting in repeat history taking or testing. Patients recounted they often had to serve as the final authority on the accuracy of information found in their EHR and perceived that providers used their knowledge as a ‘double check’ for the information in the EHR (*e.g.,* medical history, ongoing medications, or detailed reasons behind why a particular clinical decision was made). Participants reported that this redundant disclosure of their information was a source of annoyance and frustration. Others expressed some level of discomfort being the final backstop regarding the veracity of their EHR contents as they did not possess the underlying medical knowledge, did not know what information was important to healthcare providers, or could not recall all the details required.

Similarly, further risks to care were posed when inaccurate information ‘followed’ the patient through their care. Participants often reported the perpetuation of incorrect clinical information over time. Some errors, such as misdiagnoses of mental health conditions, were highlighted to be especially stigmatising when repeated across different healthcare providers or settings. For example, one participant recounted a hospital encounter where a family member was misdiagnosed as suffering from dementia upon admission. In subsequent admissions, various clinical symptoms were wrongfully attributed to being related to dementia rather than being investigated for alternative contributing aetiology. Patients wishing to correct these errors were often hampered by lengthy delays caused by various bureaucratic barriers and limited provider time to make the necessary corrections.

**Textbox 5:**
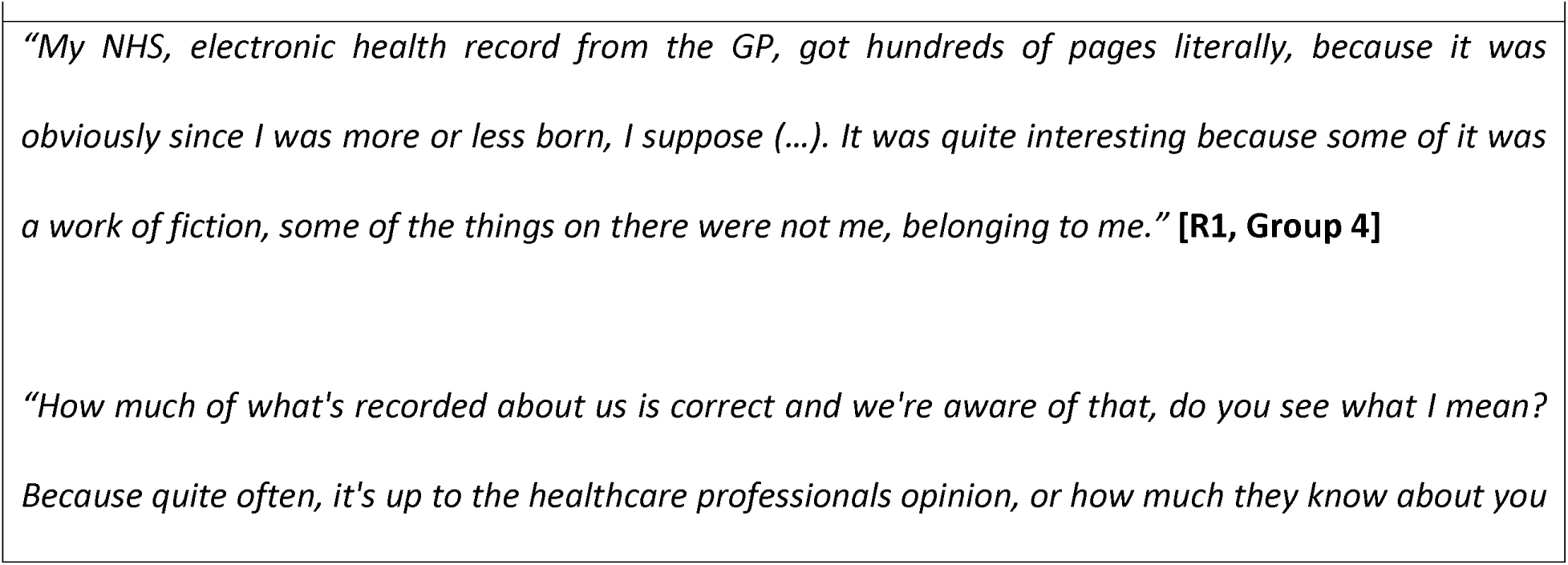
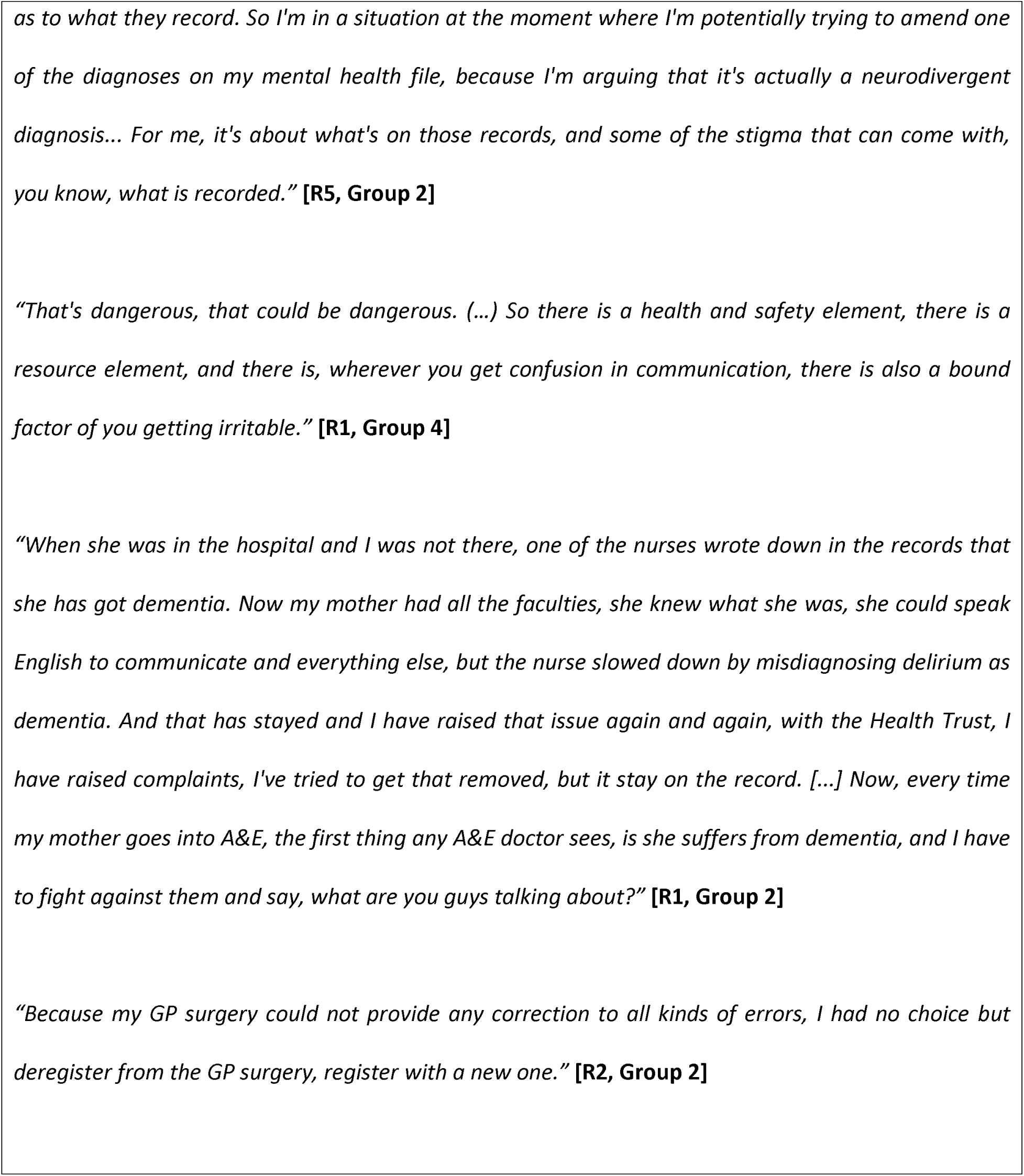
Inaccurate, incomplete, incoherent medical history.

### Negative impact on clinical decision-making

Limited EHR interoperability was perceived by participants to negatively impact clinical decision-making by healthcare providers. This was primarily due to the often inaccessible, incomplete, or inaccurate information contained within EHR on which providers had to base their decisions. No participants recounted exact instances where they experienced direct harm as a result. However, a small minority of participants were able to point to medical harm resulting from inadequate clinical information sharing reported in the recent media, and highlighted that poor interoperability at least partly contributed to those harm events.

**Textbox 6:**
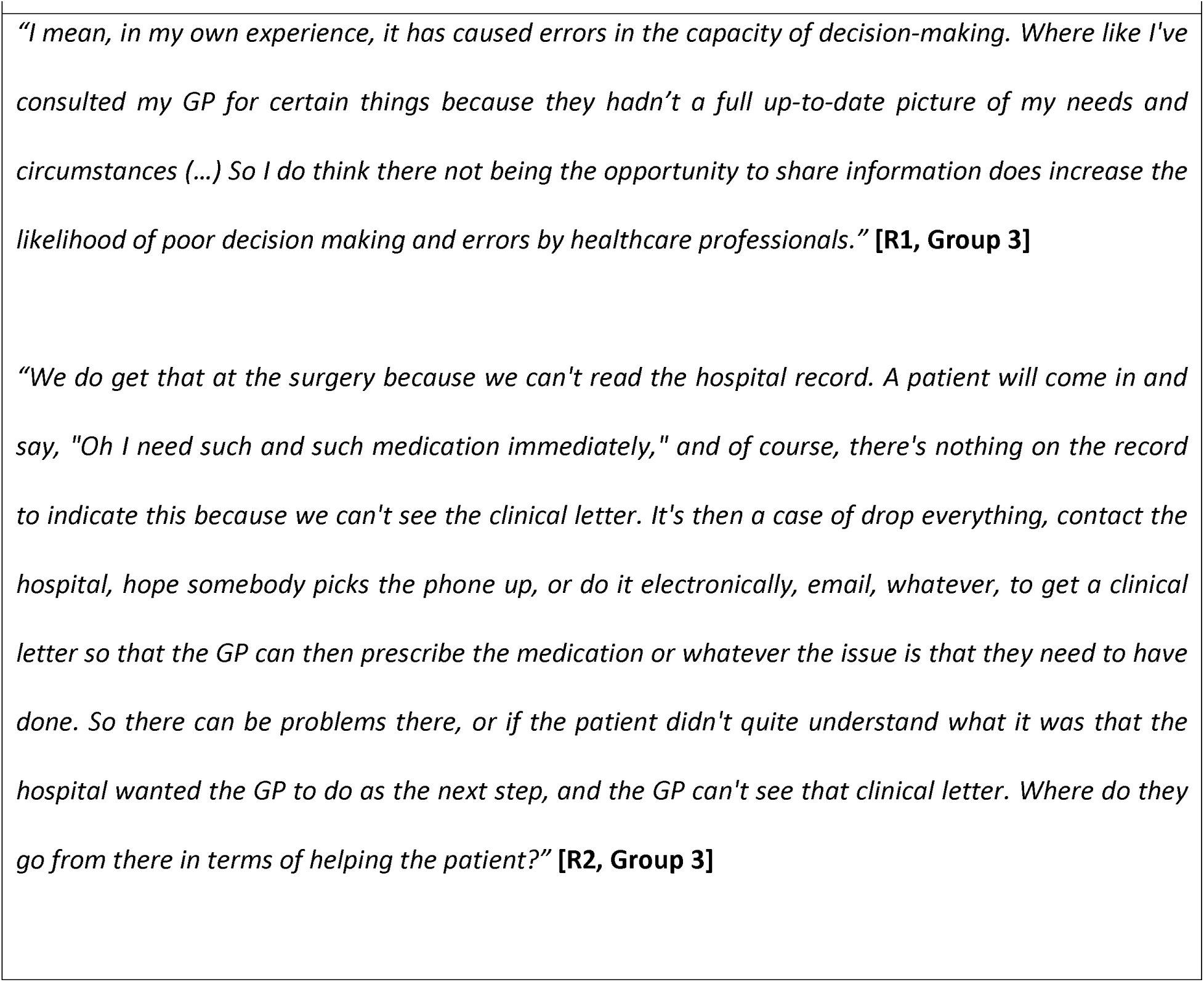
Negative impact on clinical decision-making.

### Patient-sided compromises to facilitate information sharing

Given the current lack of interoperability, participants reported that they frequently had to be the primary means of facilitating information sharing between various healthcare providers involved in their care. Inaccessible documents (*e.g.,* discharge summaries) on the EHR meant that patients frequently stepped into the role of the ‘go between’ in order to convey their care plan to their GPs as directed by hospital-based healthcare providers post discharge. In other instances, this also prompted patients to be more reliant upon physical documents (*e.g.,* GP letters, medication lists, vaccination records) as their preferred and trusted means of conveying information.

**Textbox 8:**
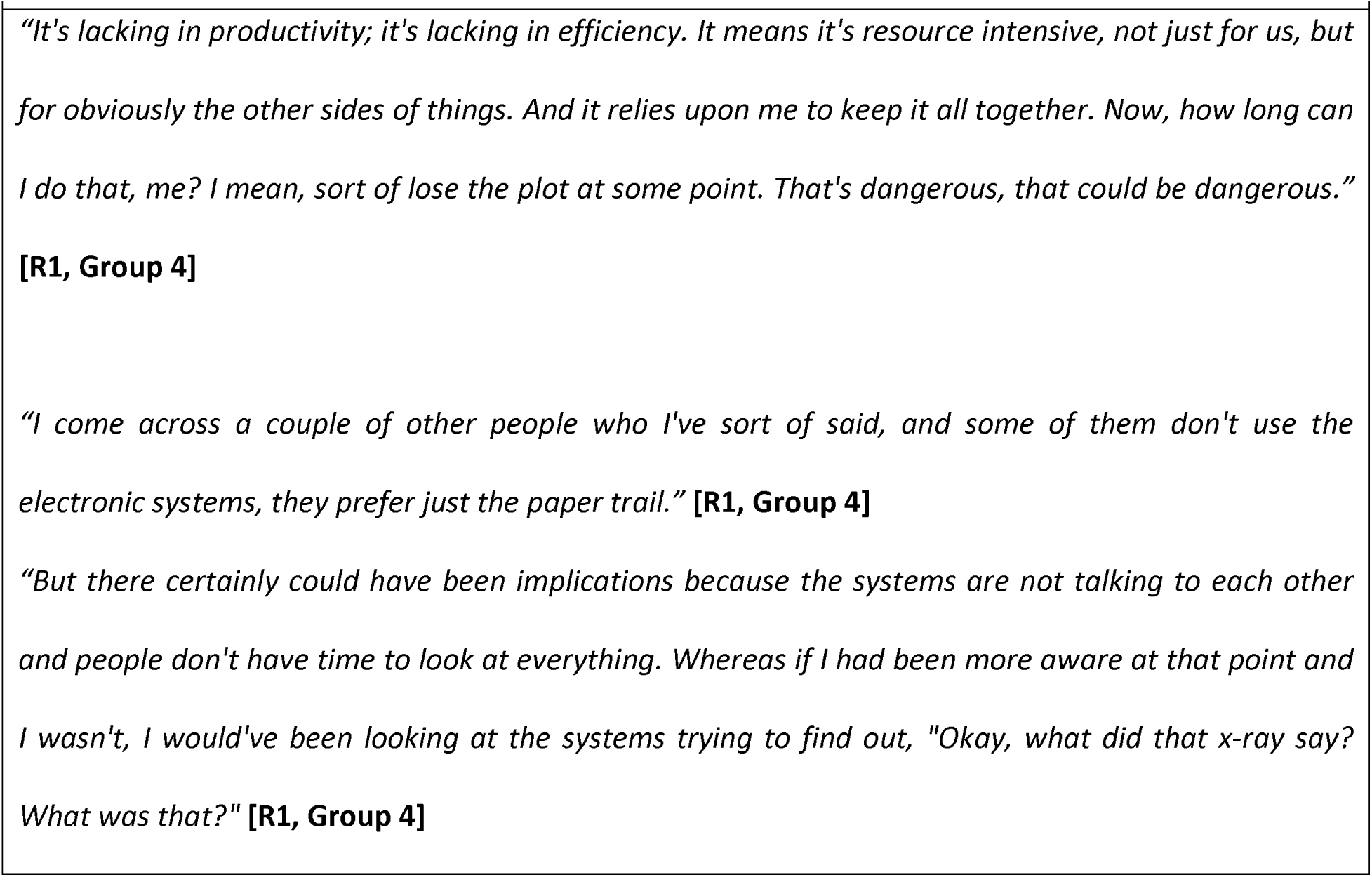
Patient-sided compromises and workarounds.

### Potential solutions to the EHR interoperability issue proposed from patients’ perspectives

#### One centralised system or application

Participants reported poor experiences trying to manage the current collection of mobile apps and online portals. Often, information in one application would conflict with that found in another and patients reported not having a way to correct it. There was near unanimous recommendation from participants for future EHR improvement efforts to be spent on having one, harmonised, interoperable approach to EHR systems in use across the NHS where their clinical data can be easily accessed, updated, and shared. However, many expressed doubts about if, how, and when this could be accomplished. Participants were aware of previous failed attempts at developing a centralised system (*i.e.,* NPfIT), but were unable to point to specific learning points which can improve a renewed attempt at a similar endeavour.

**Textbox 9:**
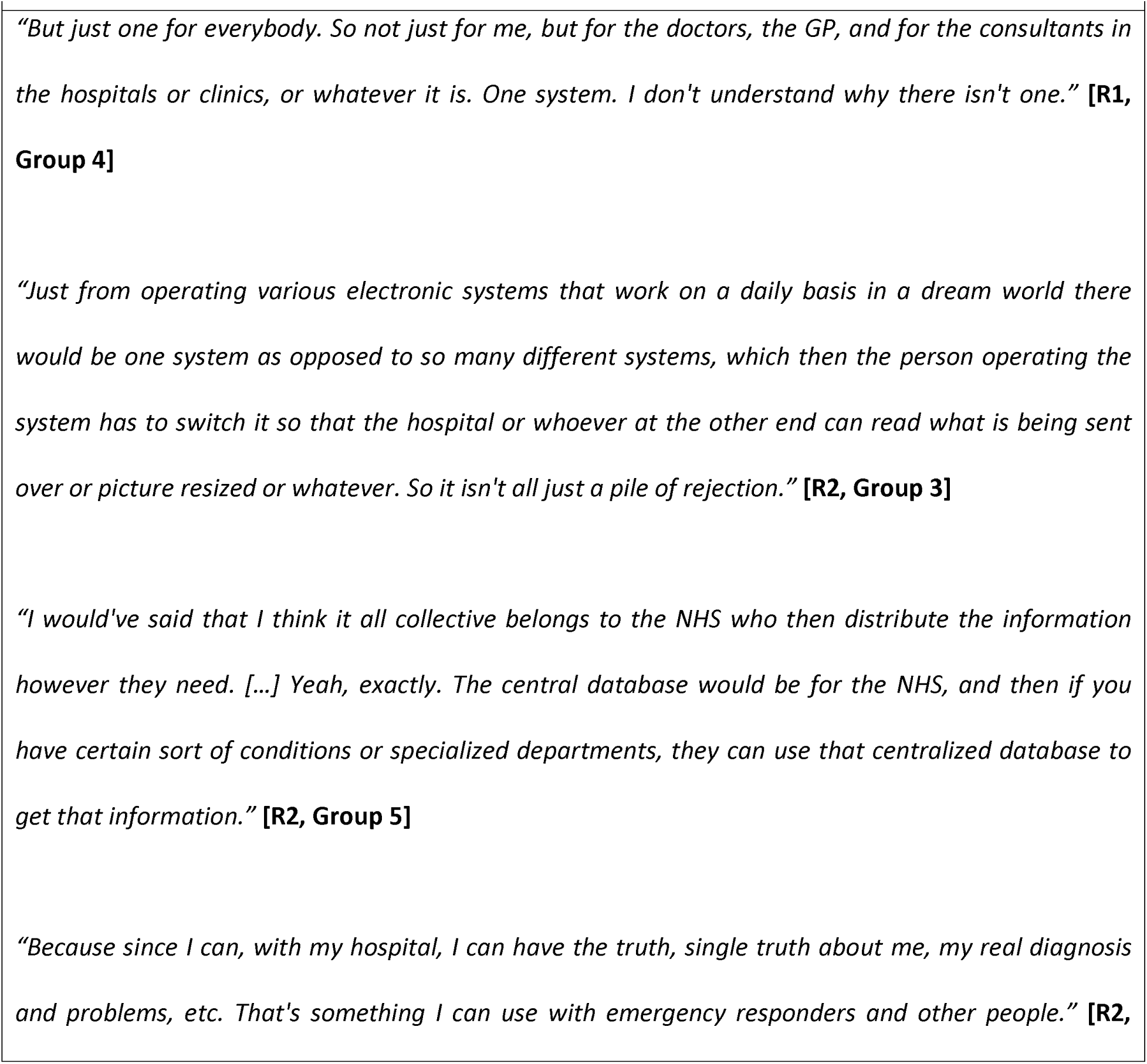
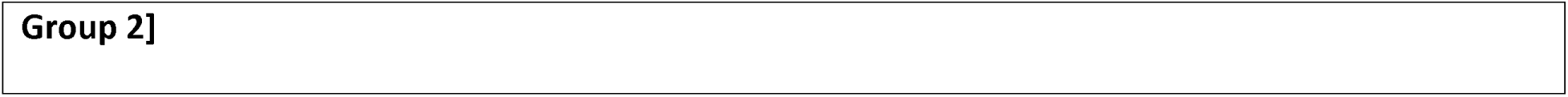
One centralised system or application.

### Strengthening data security, confidentiality, and privacy protection

Participants were weary of the elevated risks of data breaches associated with increased EHR interoperability, particularly surrounding how sensitive clinical information (*e.g.,* diagnoses regarding mental health, sexually transmitted diseases) may be mishandled with highly stigmatising ramifications. Most were clear that the greater dissemination of clinical data must not be commercialised or used to identify financial or political information. Participants emphasized the need for strengthened data protection measures in parallel with interoperability improvement efforts.

**Textbox 10:**
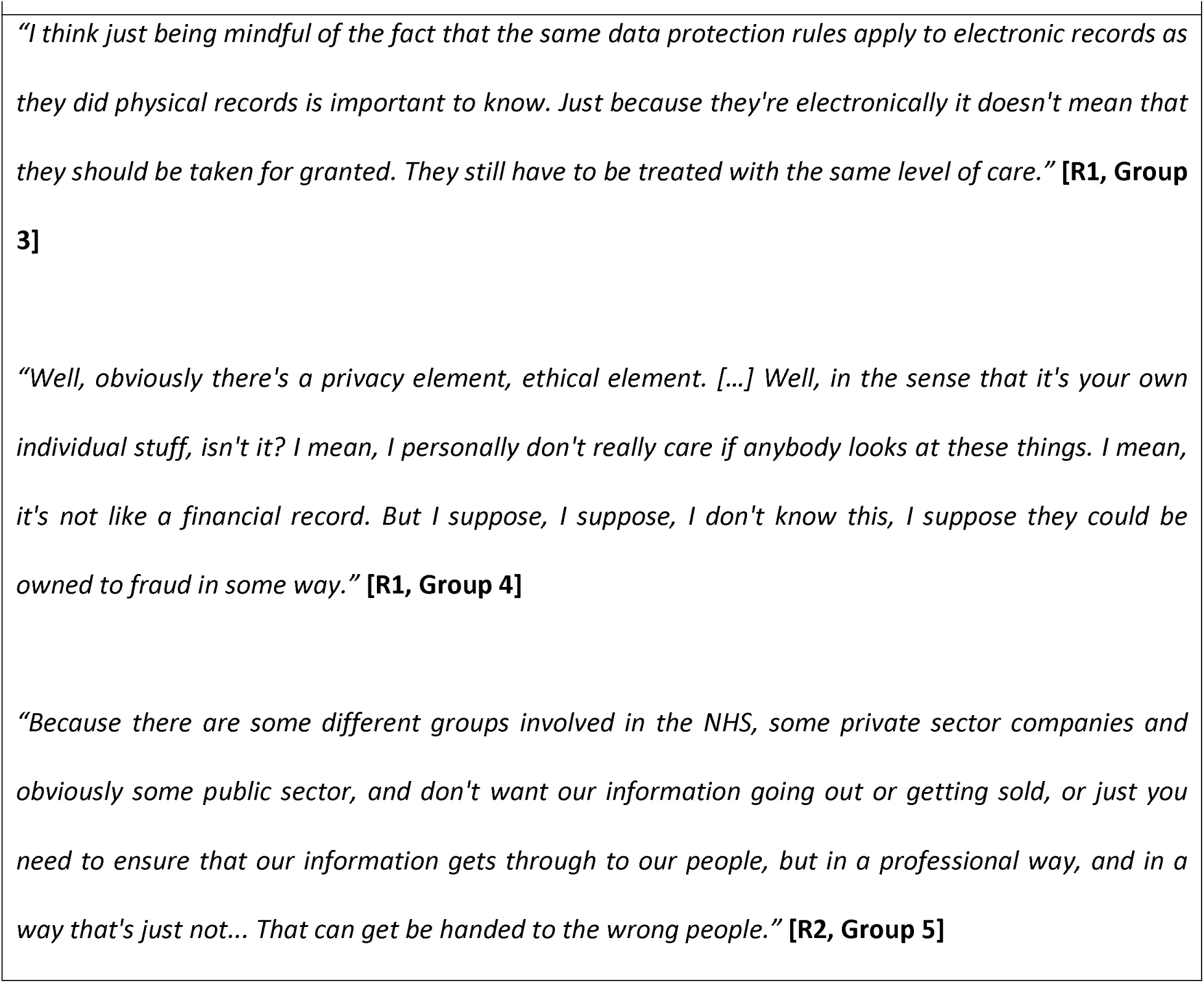
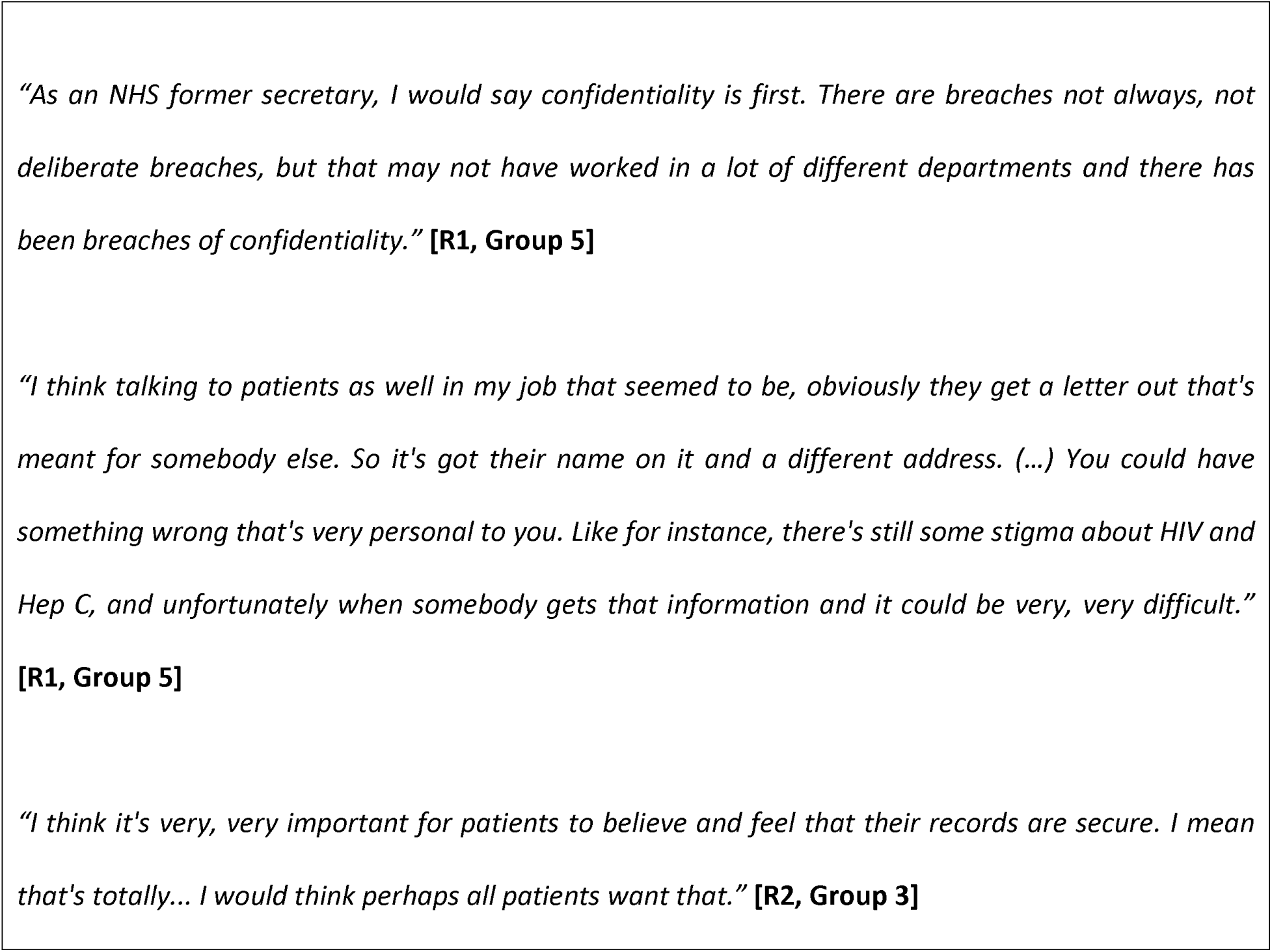
Strengthening data security, confidentiality, and privacy protection.

### Supporting greater patient involvement in EHR data curation

Greater support for patient involvement in their own EHR data to improve its overall accuracy, completeness, and quality was expressed by almost all study participants. Patients noted that interoperability between EHR was only valuable so long as data within it was accurate. At present, their health information within EHR is often inconsistent and incomplete, thus limiting the overall user experience for patients even when interoperable systems were made available. By contributing to the curation of own health data, patients thought that they can indirectly help efforts to hasten interoperability by contributing to the quality of the data in the records. Patients believed they are well-suited to play a constructive role in achieving that aim, especially when clinicians simply do not have the resources or time to do so.

However, views varied greatly surrounding the extent to which patients should be able to directly intervene in their own clinical data found in EHR. Most participants reported that while they would appreciate having greater access, the final authority on the information documented should remain with healthcare providers, ideally their GPs. Some participants acknowledged that asking their GPs to do so is unlikely to be feasible given their workloads and proposed a more centralised NHS entity dedicated to managing EHR data on behalf of both providers and patients. Patients believed their own contribution should come in the form of a partnership by flagging errors themselves so that they are quickly addressed while leaving more technical, clinical information curation with healthcare providers.

Divergent views included participants who believed patients would not care to be involved so long as the information was well-maintained. A small minority expressed concerns that patient involvement may be detrimental in cases when over-enthusiastic patients scrutinising their own records may generate increased workloads for healthcare staff needing to address every concern raised.

**Textbox 11:**
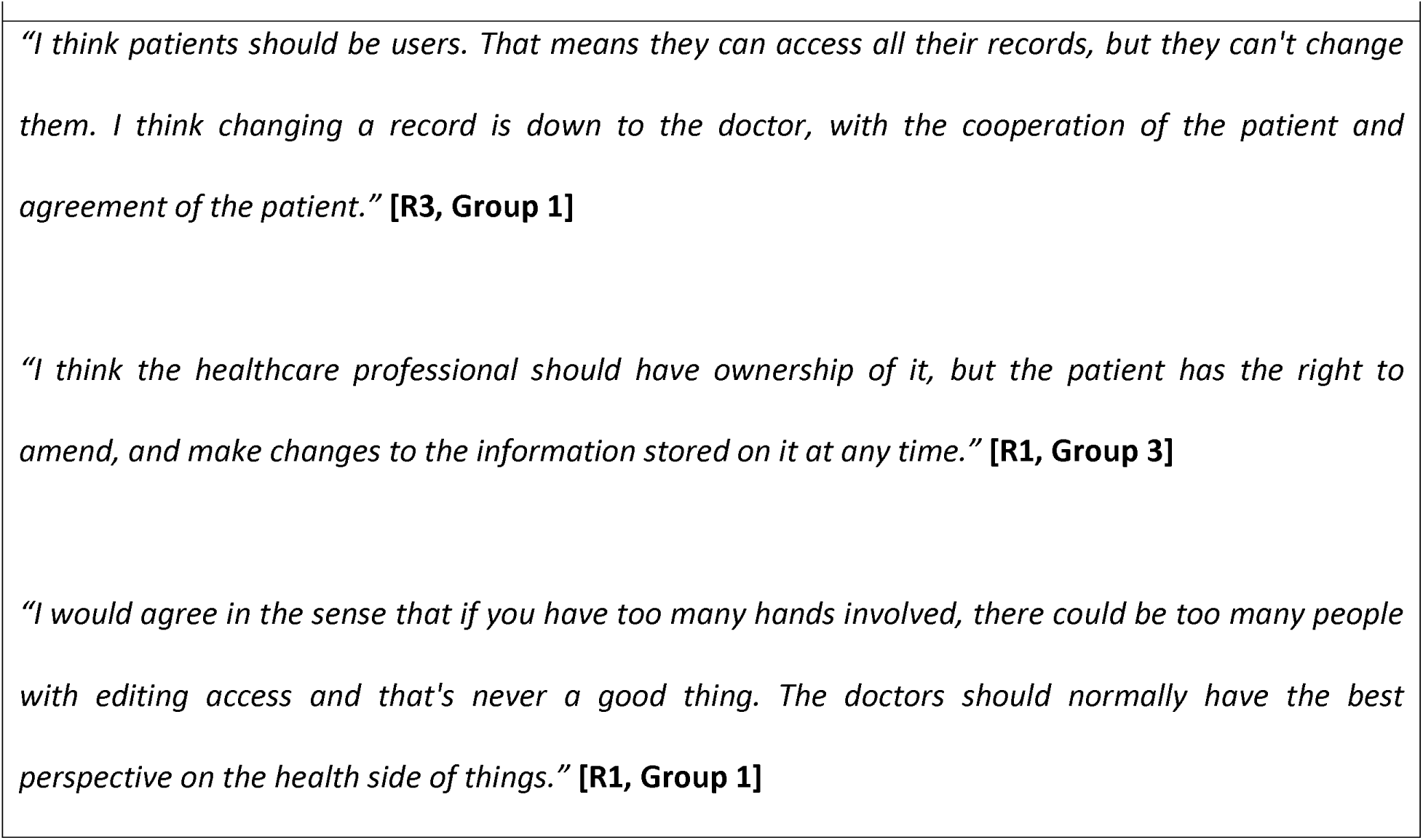
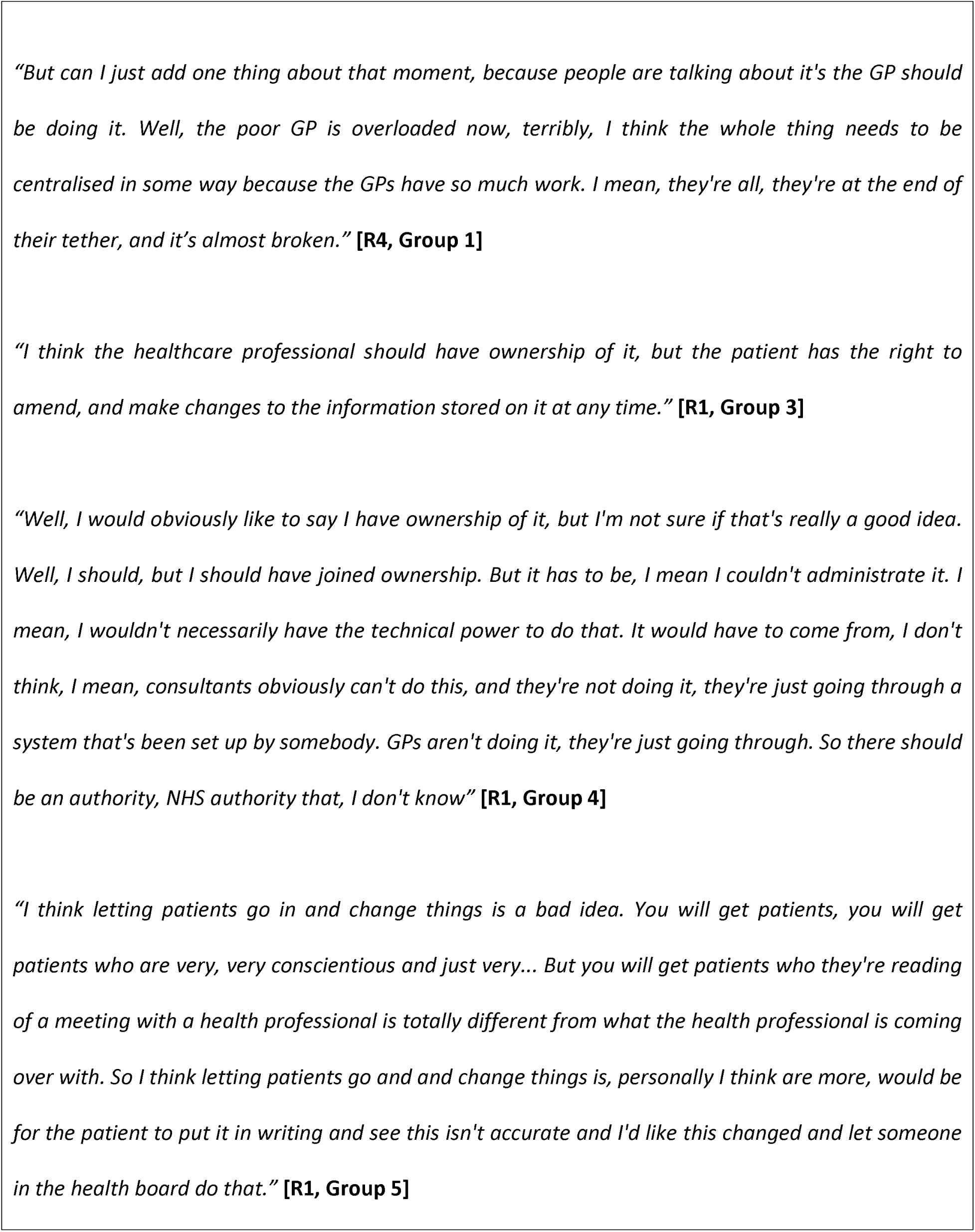
Supporting greater patient involvement in EHR data curation.

### Prioritising more HIT training for NHS staff

Greater investment in HIT training for NHS staff, both frontline healthcare workers as well as administrators involved in the maintenance and procurement of EHR systems, was a point for improvement identified by patients. More standardised training and familiarisation with the various EHR platforms available, their functions, and how to perform common procedures (*e.g.,* inputting GP letters, filing a scan/lab result), was perceived to facilitate advancement of user-level interoperability. Participants believed that training for administrators would also be beneficial to ensure that EHR systems procured are aligned with the needs of healthcare providers.

**Textbox 12:**
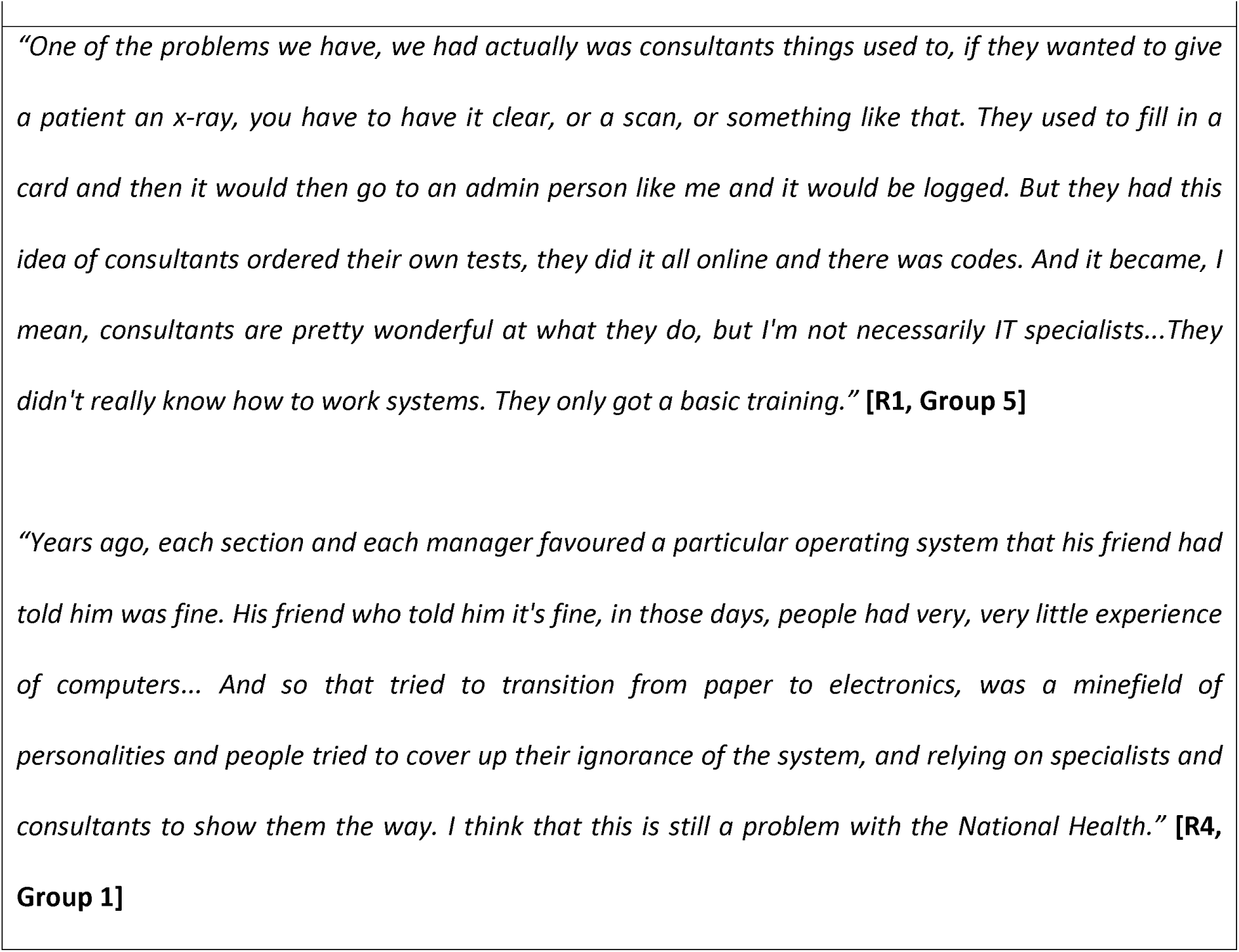
Prioritising more HIT training for NHS staff.

### Greater involvement of allied health services and social care

Participants reported that greater involvement of allied health professionals would not only help with the comprehensiveness of the stored data but may also serve as a backstop to doctors in spotting errors, particularly of non-medical issues such as social care needs. As many patients are attended to by a range of healthcare staff, participants expressed the desire that future improvements to EHR and interoperability should work towards a more holistic medical record that is better representative of their overall health and well-being.

**Textbox 13:**
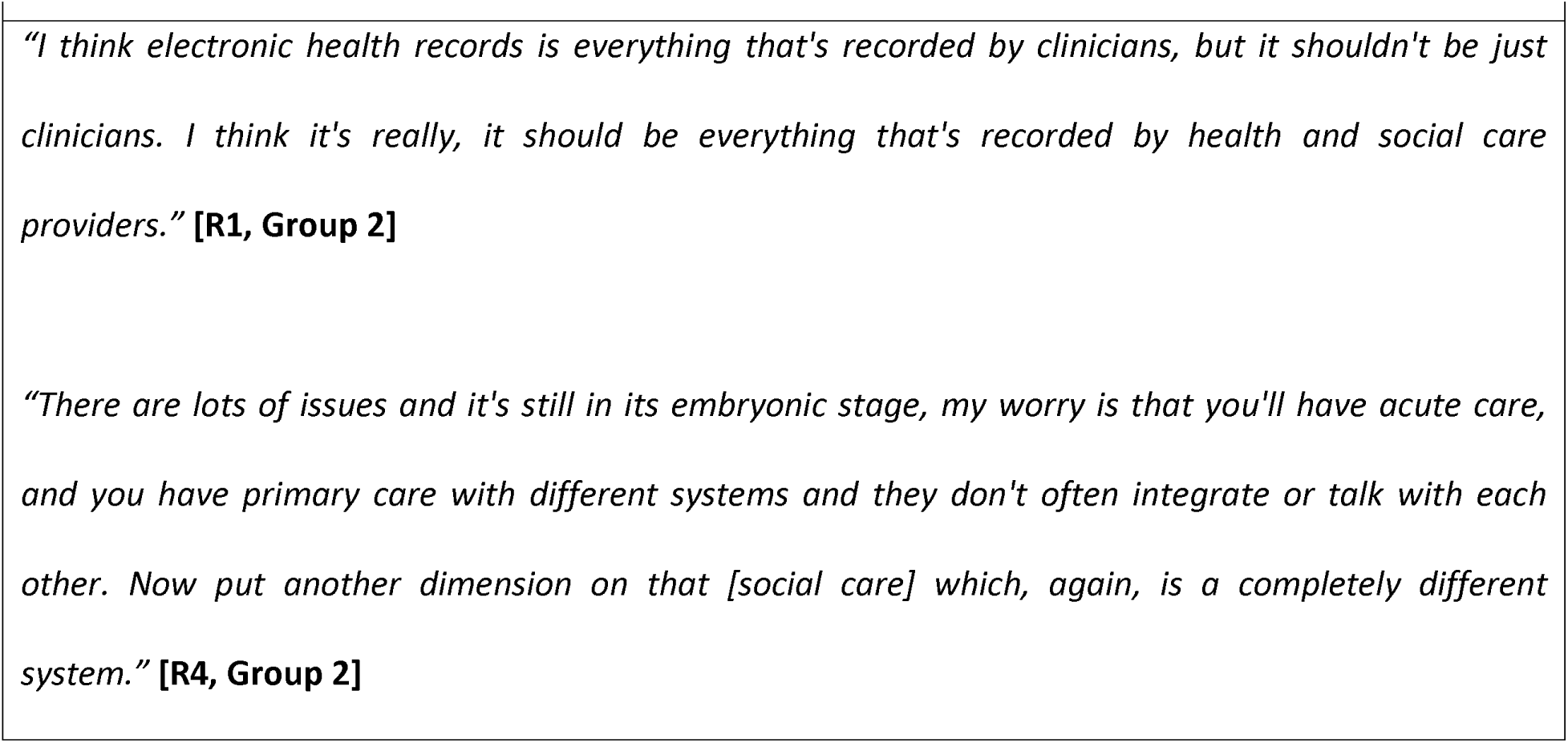
Greater involvement of allied health services and social care.

## Discussion

### Summary of Main Findings

Interoperability was largely an unfamiliar concept to patients and caregivers. Once a brief definition was provided however, most participants recognised its necessity and potential benefits. Participants were able to highlight how the lack of EHR interoperability negatively impacted their care, predominantly as a source of notable inconvenience for themselves and their healthcare providers.

Limited interoperability has reportedly contributed to inaccurate, incomplete, or incoherent medical histories within EHR which were often difficult to amend, thereby leading to the unencumbered perpetuation of errors. Patients recounted that this has negatively impacted clinical decision-making. Consequences ranged from repeat history taking or investigations, to more serious ones such as misdiagnoses of potentially stigmatising conditions. Patients regularly had to serve as the main means to ensure information was shared appropriately. This often included carrying paper-based documentation when travelling between providers, verbally conveying care plans, and acting as the final backstop for providers to verify clinical information found within EHR.

Participants proposed several solutions or factors which should be considered in any future attempts to address poor EHR interoperability. Most participants conveyed a desire for *‘one centralised system’*. As improving interoperability will result in greater amounts of clinical information being shared, participants stressed the importance of a corresponding increase in data and privacy protection measures. There was an overarching sentiment that efforts to improve provider-to-provider information sharing should also include parallel efforts to improve information sharing with patients. Patient participants perceived that increased interoperability with their involvement would attenuate the current risks of information sharing, namely sharing of inaccurate information between different providers. More robust training for NHS staff to foster better HIT competency and greater involvement of allied health services and social care in the upkeep of EHR content were areas in which participants believe further efforts should be made to improve overall EHR interoperability.

### Strengths and Limitations

This study contributes to the growing body of evidence investigating how patient perspectives can be used to inform future directions for key healthcare challenges. The topic guide was informed by existing literature and was piloted with two patient research groups before use. The focus groups were conducted online, which allowed patients and caregivers from across the UK to participate and thus broadened the range of perspectives included.

However, our findings must also be taken in the context of certain limitations. Firstly, the relatively small number of participants, mostly of elderly age, limits the range of views and perspectives discussed in our focus groups. There is also some level of self-selection bias as participants are likely to be patients who feel more strongly about their views or are more able or inclined to participate.

The inclusion of only adult patients with chronic conditions also neglects exploration of the views of other patient groups, such as parents and children. The inclusion of only patients who were able to participate using video conferencing introduced selection bias for those who are more digitally literate, and likely from socio-economic backgrounds who could afford the technology necessary. Lastly, the restriction of participants to only those able to converse in English likely contributed to excluding those less proficient, or unable to speak the language.

### Comparison with Prior Work

Few studies have investigated specifically EHR interoperability and its practical implications on patient care from their perspective (26).

*Kelly et al.,* summarised how current EHR interoperability levels contributed to suboptimal care for dialysis patients in the US (27). Specifically, patient perceptions of HIE were found to be dependent on the perceived risks and benefits to their care resulting from increased clinical data sharing, which were based on bespoke lived experiences. For example, patients from ethnic minority backgrounds were found to have greater concerns regarding the sharing of their clinical information through HIEs. The authors also pointed to potential solutions to the patients identified to mitigate these concerns, many of which align with those reported in our own study including greater patient engagement and increasing opportunities for patient feedback longitudinally (27).

In 2016, *Legler et al.,* conducted a survey study which explored patient satisfaction levels and their perception of their healthcare provider’s knowledge of their medical history after the introduction of a centralised portal (26). While the portal did not allow for the manipulation or transfer of patient health data stored across the US VA, Department of Defence, and community providers, the authors nonetheless found that merely using it to view its contents resulted in patients being 14% more likely to report higher levels of satisfaction with their healthcare providers as they appeared more knowledgeable about their medical history (26). This was especially noticeable for patients with long-standing relationships with their healthcare providers (26). However, the authors also noted that providers were more likely to use the portal with newer patients and thus be the ones more likely to reap the greatest clinical benefits compared to patients with existing relationships (26).

A recent publication by *Hussein et al.,* investigated patient-generated health data (PGHD) interoperability from three distinct perspectives (technological, clinical, and users) concerning the proposed DH-Convenor platform, an initiative to support the collection, organisation, and interoperability of PGHD in the Austrian national health system (28). Many of the overarching opportunities and barriers identified mirror the views expressed by our study participants. For example, the authors found that patients were largely accepting of the clinical necessity for PGHD interoperability and its purported benefits. However, participants clearly opposed the sharing of socioeconomic data and any attempts to use patient data for commercial or political purposes (28).

A qualitative study by *Sanyer et al.,* examined patient perspectives on clinical information sharing via EHR in team-based care (29). Though authors noted that patients recognised the value of centralising their clinical information, concerns were raised towards documentation inaccuracies and the associated difficulties in resolving them (29). Patients acknowledged that the ability to share health information amongst members of their care team was important, but also voiced apprehensions regarding data security, who should have access to their information, and the scope of information shared (29). This was especially poignant for sensitive clinical information such as mental health diagnoses. Aside from the sharing of ‘basic’ information (*e.g.,* name, date of birth) with all members of the care team, patients were only willing to share more sensitive or personal information found in their EHR with their immediate healthcare provider or someone the patient already had a well-established relationship with (*i.e.,* GP) (29).

### Implications for Policy, Practice, and Further Research

The past two decades of EHR use have demonstrated the importance of greater data sharing and the necessity of interoperability in the HIT infrastructure of modern healthcare systems. As demonstrated in our study, the need for a more coordinated, interoperable EHR system is evident. Despite the priority often being placed on tailoring such systems to the requirements of healthcare providers, our recent experiences, especially throughout the pandemic, have made apparent the urgency with which future EHR systems and the policies surrounding their use, must better accommodate the needs and expectations of patients. Tackling the lack of interoperability will necessitate fundamental changes ranging from the types of data that are stored and how it is shared, to addressing more systemic questions such as aligning information governance with evolving patient preferences. A renewed attempt at realising some form of a centralised EHR system should be revisited, especially given that the recognition of its need is apparent even amongst patients.

As shown in our findings, the idea of increased EHR interoperability is synonymous with greater access to patient data for more stakeholders, including for patients themselves. HIT policies and procurement contracts must encourage vendors to incorporate this into their design of future EHR systems from the outset and ensure that as information sharing is enhanced for providers, it is also made more robust for patients. As illustrated by our participants and supported by findings from other similar studies, the greater sharing of clinical information possible through better interoperability is contingent upon efforts to strengthen data security and privacy protections (28–30).

Future research should seek to quantify any improvements through the implementation of new EHR interoperability interventions. Outcome measures of interest may include the accuracy of stored clinical information, patient safety incidents, user satisfaction, or time spent reconciling patient information. These efforts would help demonstrate the value of expanding EHR interoperability and serve to corroborate the findings found in patient-centric qualitative studies such as our own. Likewise, additional qualitative studies focussing on underrepresented ethnic minority groups, non-English speaking communities, and different target patient demographics (*e.g.,* paediatrics), would also be useful in ensuring future efforts at improving EHR interoperability are inclusive of their needs.

## Conclusion

Our study has demonstrated that patients and caregivers are keenly aware of how the current state of NHS EHR interoperability affects the safe and efficient delivery of their care. Their perspectives offer policymakers and health information technologists valuable insight into the impact current levels of EHR interoperability have on clinical care for patients with complex conditions and illustrates patient-derived solutions for enhanced interoperability.

In its current form, EHR interoperability remains lacklustre in fulfilling the needs and expectations of patients, often causing inconvenience for users, contributing to inaccurate or incomplete medical histories, impairing clinical decision-making for healthcare providers, and requiring clumsy workarounds from patients themselves to mitigate its deficiencies. Only by comprehensively addressing these issues raised by patients would health systems be able to realise the purported benefits of EHR with greater interoperability.

## Declarations

### Consent for Publication

All participants provided written informed consent for anonymised quotes from their focus groups to be used in the publication of this study.

### Availability of Data and Materials

Data is available upon reasonable request to the corresponding author.

## Ethical approval

Overall ethical approval for this study was granted by the Imperial College Research Ethics Committee (ICREC) (Reference No. 22IC7425). This is a dedicated ethics oversight body at Imperial College London for all health-related research involving human participants. All participants provided written informed consent prior to participating in the study and consented to their focus group sessions being recorded.

## Author contributions

EL and ALN conceived of and designed the study. EL and OL conducted the focus groups. EL and OL analysed and interpreted the resultant transcripts. EL drafted the initial manuscript. EL, OL, and ALN contributed to writing the manuscript. EL, ALN, OL, JC, HA, and AD were all contributors to editing the manuscript. All authors have read and approved the contents of the final manuscript.

## Data Availability

Data is available upon reasonable request to the corresponding author.

## Abbreviations

COREQ: Consolidated criteria for Reporting Qualitative research
EHR: electronic health records
GP: general practitioners
HIE: health information exchange
HIT: health information technologies
JLV: Joint Legacy Viewer
NHS: National Health Service
PGHD: patient-generated health data
PHR: personal health records

## Appendix

## Appendix 1

[Insert focus group topic guide]

## Appendix 2

[Insert ethics approval form]

