## Appendix 1 for "Patient and caregiver perceptions of electronic health records interoperability in the NHS and its impact on care quality: A focus group study"

**Document version: 1.0**
**Document date: 26 June 2023**
**ICREC ID: 22IC7425**

### Project title: Patient and caregiver perceptions of electronic health records interoperability and impact on care quality

**Going over the Participant Information Sheet:**

- Overview and purpose of study
- Aims of the focus and expected duration
- Who is involved?
- Why participant's involvement is important, advantages and disadvantages
- What will happen to the results of this study?
- Any questions?

**Summary of study objectives:** *Before I ask you further questions, here is what we will be covering in today's discussion:*

1. To explore what are patients'/caregivers' knowledge, understanding, and expectations of concepts such as: electronic health records and interoperability.
2. To explore what were some practical patient safety implications limited interoperable EHRs had on patients' care.
3. To investigate any incidents in the past where limited EHR interoperability resulted in suboptimal/unsafe care and how it arose.
4. To explore any potential solutions to the EHR interoperability issue proposed from the patients' perspective.

| **Aim(s)** | **Topics & Prompts** | **Examples/Clarifications** |
|---|---|---|
| To explore what are patients'/caregivers' knowledge, understanding, and expectations of concepts such as: electronic health records and interoperability. | **1. In your opinion, what is an electronic health record?**<br><br>**2. Interoperability – are you aware of what this is? What does this mean to you?** | • EHRs are defined as: *computer software that physicians use to track all aspects of patient care. Typically, this broader term also encompasses the practice management functions of billing, scheduling, etc.*<br><br>• INTEROPERABILITY is defined as: *the ability of health information systems to work together within and across organisation boundaries to advance effective delivery of healthcare for individuals and communities.* |
| To explore what were some practical patient safety implications limited interoperable EHRs had on patients' care. | **1. How do you feel EHRs typically affect your experience receiving care from the NHS?**<br><br>**2. Do you think EHRs and interoperability are important to how patients engage with healthcare providers?**<br><br>**3. Do you think patients want more access and control over their own medical records through EHR interoperability?**<br><br>**4. Does EHR and interoperability affect your daily life outside of your immediate interactions with the healthcare system or doctor? If so, how?** | • Ask for examples whenever possible. |

| | |
|---|---|
| To investigate any incidents in the past where limited EHR interoperability resulted in suboptimal/unsafe care and how it arose. | **1. Based on your experiences of receiving care at NHS healthcare facilities, tell us about any instances where something went wrong or nearly went wrong, or when you felt the care was unsafe because of EHRs or interoperability-related issues.** |
| To explore any potential solutions to the EHR interoperability issue proposed from the patients' perspective. | **1. What kind of changes/solutions do you think are needed to improve patient safety when it comes to the use of technologies such as EHRs and the accompanying problem of interoperability?**<br><br>**2. What are some barriers or facilitators do you foresee in bringing about these changes?** |

**Closing:** Is there anything else that you think is important about using electronic health records, interoperability, and patient safety that we have not talked about?

- Summarize covered research domains/questions.
- Any other questions or concerns?
- Ensure participant has copies of participant information sheet and consent forms; provide any additional information as needed
- Thank participants for their input and engagement in the discussion.
