## Appendix 2 for "Patient and caregiver perceptions of electronic health records interoperability in the NHS and its impact on care quality: A focus group study"

Imperial College London

**Imperial College Research Ethics Committee**
Imperial College London
Room 221
Medical School Building
St Marys Campus
London
W2 1PG

22/02/2022

Dear Dr. Ana Luisa Neves,

**Study Title:** Patient and caregiver perceptions of electronic health records interoperability and its impact on care quality

**ICREC Reference number: 22IC7425**

The above study was approved by your Head of Department on 24/12/21 and by the Research Governance and Integrity Team (RGIT) on 22/02/22.

Under the Imperial College Research Ethics Committee process, a study that has been reviewed by the Research Governance and Integrity Team (RGIT) and Head of Division/Department, where no significant ethical issues have been identified in the protocol or ethics application, can be granted RGIT approval without requiring it to go to full committee.

**Documents**
The documents reviewed were:

- Application form v1.3 21/02/2022
- Protocol v1 10/12/2021
- Participant Information Sheet v1.2 14/02/2022
- Consent form V1 13/12/2021
- Invitation E-mail v1 22/02/2022
- Topic Guide v1 14/02/2022

Yours sincerely

Ruth Nicholson Digitally signed by Ruth Nicholson
Date: 2022.02.22 15:29:46 Z

Ruth Nicholson
Head of Research Governance and Integrity
Imperial College London
